# Early surveillance and public health emergency disposal measures between novel coronavirus disease 2019 and avian influenza in China: a case-comparison study

**DOI:** 10.1101/2020.03.29.20046490

**Authors:** Tiantian Zhang, Wenming Shi, Ying Wang, Ge Bai, Ruiming Dai, Qian Wang, Li Luo

**Affiliations:** School of Public Health, Fudan University, Shanghai 200032, China; Key Laboratory of Public Health Safety of the Ministry of Education and Key Laboratory of Health Technology Assessment of the Ministry of Health, Fudan University, Shanghai 200032, China

**Keywords:** COVID-19, Emerging infectious diseases, H7N9, Emergency management, China

## Abstract

**Background:** The novel coronavirus disease 2019 (COVID-19) outbreak is spreading rapidly throughout China and the world. Hence, early surveillance and public health emergency disposal are considered crucial to curb this emerging infectious disease. However, studies that investigated the early surveillance and public health emergency disposal for the prevention and control of the COVID-19 outbreak in China are relatively few. We aimed to compare the strengths and weaknesses of early surveillance and public health emergency disposal for prevention and control between COVID-19 and H7N9 avian influenza, which was commended by the international community, in China.

**Methods:** A case-comparison study was conducted using a set of six key time nodes to form a reference framework for evaluating early surveillance and public health emergency disposal between H7N9 avian influenza (2013) in Shanghai, China and COVID-19 in Wuhan, China.

**Findings:** A report to the local Center for Disease Control and Prevention, China, for the first hospitalized patient was sent after 6 and 20 days for H7N9 avian influenza and COVID-19, respectively. In contrast, the pathogen was identified faster in the case of COVID-19 than in the case of H7N9 avian influenza (12 days vs. 31 days). The government response regarding COVID-19 was 10 days later than that regarding avian influenza. The entire process of early surveillance and public health emergency disposal lasted 5 days longer in COVID-19 than in H7N9 avian influenza (46 days vs. 41 days).

**Conclusions:** The identification of the unknown pathogen improved in China between the outbreaks of avian influenza and COVID-19. The longer emergency disposal period in the case of COVID-19 could be attributed to the government’s slower response to the epidemic. Improving public health emergency management could lessen the adverse social effects of emerging infectious diseases and public health crisis in the future.

## Introduction

In the past 20 years, China has experienced several public health crises due to infectious disease outbreaks, such as severe acute respiratory syndrome in 2003, H1N1 swine influenza in 2009, and H7N9 avian influenza in 2013, seriously impacting health, economy, and global security. (1-3) These outbreaks challenged the health emergency management in several countries, especially developing countries, including China. (4, 5) In late December 2019, the novel coronavirus disease 2019 (COVID-19) emerged in Wuhan City, China, and rapidly spread throughout China and the world. (6) Prior to March 5, 2020, the Chinese government reported 80,409 confirmed cases and 3,012 fatalities due to COVID-19. (7)

COVID-19 and H7N9 avian influenza are emerging infectious diseases that share similar characteristics (Table 1), such as probable development of severe respiratory diseases and susceptibility regardless of age. However, the socioeconomic losses were higher in COVID-19 than in H7N9 avian influenza. An effective public health emergency management reduces the adverse impact of emerging infectious diseases (8). This management relies on the early surveillance and timely information dissemination available in a given period. (9) The following three key responses are often analyzed to evaluate the efficiency of public health emergency disposal: (1) time taken by the hospital to report an emerging infectious disease, (2) time taken to identify the pathogen, and (3) time taken by the government to respond. (10-12)The World Health Organization declared a Public Health Emergency of International Concern on January 30, 2020. (13) Since then, China established and strengthened the national and local surveillance systems as well as emergency responses to prevent and control the spread of COVID-19. (14) Comparing the infectious disease surveillance and public health emergency disposal between different outbreaks in China could assist in improved public health strategies and decision-making by the government to prevent and control future epidemics, both in China and the world. To the best of our knowledge, few studies have been conducted to investigate the early disease surveillance and public health emergency disposal between other epidemics and COVID-19 in China.

**Table 1.** Characteristics of the H7N9 avian influenza and coronavirus disease 2019 in China.

| Characteristics | H7N9 | COVID-19 |
| --- | --- | --- |
| Country of origin | China | China |
| First case in China | February 2013 in Shanghai | December 2019 in Wuhan |
| Viral genome | Negative segmented RNA | Positive single-stranded RNA |
| Pathogen identification | CDC, China; March 29, 2013 | CDC, China; January 7, 2020 |
| Human-to-human transmission | Limited | High |
| Genesis/source | Domestic poultry | Unclear (so far) |
| Method of diagnosis in China | Real-time PCR | Real-time PCR |
| Vaccines in China | Not yet available | Not yet available |

In the present study, we aimed to compare the key time nodes of early surveillance and public health emergency disposal to prevent and control between COVID-19 and H7N9 avian influenza.

## Methods

### Data collection

Data regarding the public health emergency disposal of the novel COVID-19 in Wuhan City, China, were obtained from published literature, secondary statistical data, WHO reports, (3) official websites (e.g., National Health Commission of the People’s Republic of China [http://en.nhc.gov.cn/], Chinese Center for Disease Control and Prevention [CDC] [http://www.chinacdc.cn/en/], Health Commission of Hubei Province, and Wuhan Municipal Health Commission), and credible media reports in China (CCTV, People’s Daily, CBN, YiMagazine). Data regarding H7N9 avian influenza in Shanghai, China, were obtained from our published literature. (15)

### Comparative analysis

We compared the six key time nodes during the entire period from the detection of the first case to the launch of the health emergency response between COVID-19 in Wuhan City and H7N9 avian influenza in Shanghai. They were as follows: hospitalization of the first case, hospital report to the local CDC, laboratory identification of the pathogen, technical recheck of the pathogen, confirmation and notification of the pathogen, and launch of emergency disposal through the Chinese government.

We further evaluated the following three crucial periods during the public health emergency disposal of emerging infectious diseases: time taken by the hospital to report a case to the local CDC, time taken to identify the pathogen i.e., organization of the CDC laboratory to detect and recheck the pathogen, and time taken by the government to respond i.e., implementation of the emergency response once the pathogen is confirmed. Moreover, we calculated the number of days during each time node using the hospitalization time reference of the first case as the benchmark. The duration between detecting the first case and report the first death was also analyzed in the study.

## Results

The comparison of three crucial periods between COVID-19 in Wuhan City and H7N9 avian influenza in Shanghai are shown in Table 2 and Figure 1. The entire process of early surveillance and public health emergency disposal was 5 days longer in the case of COVID-19 than in the case of H7N9 avian influenza (46 days vs 41 days). The details regarding the comparative analysis using the set of six key time nodes and three crucial time periods are as follows.

**Table 2.**
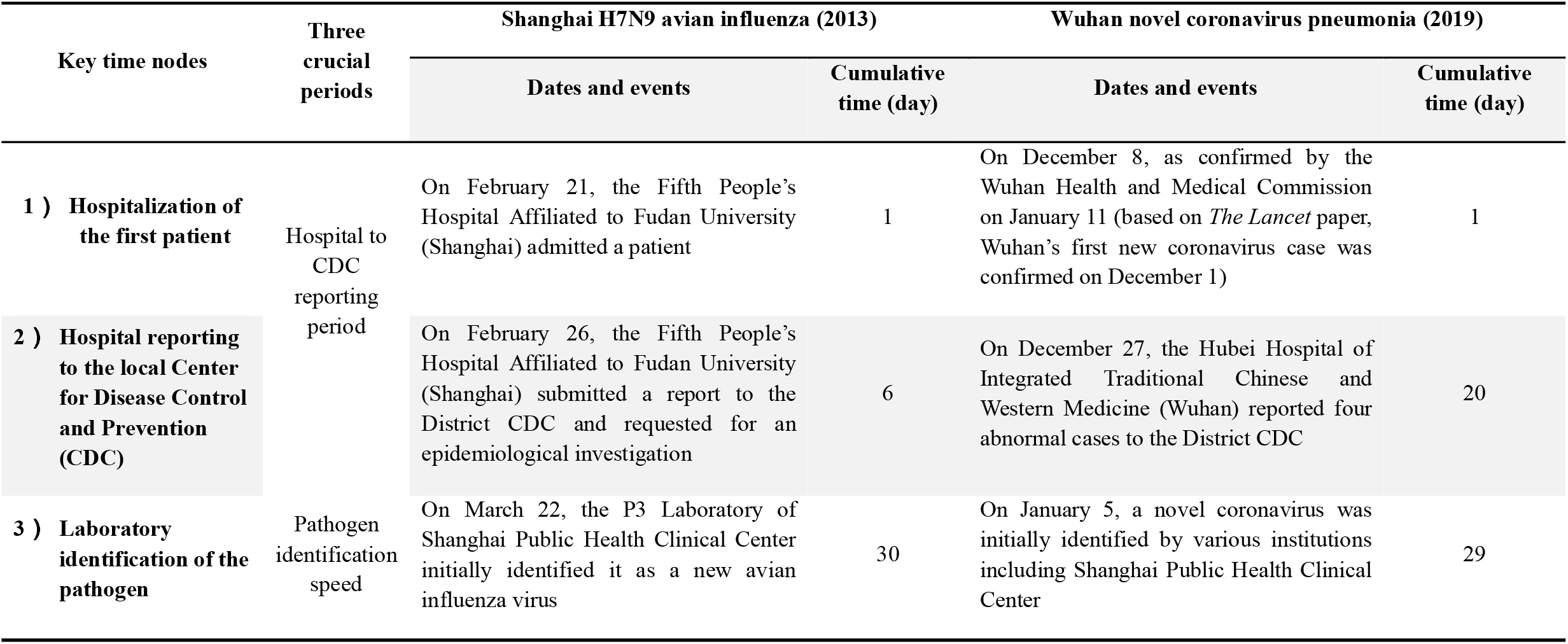
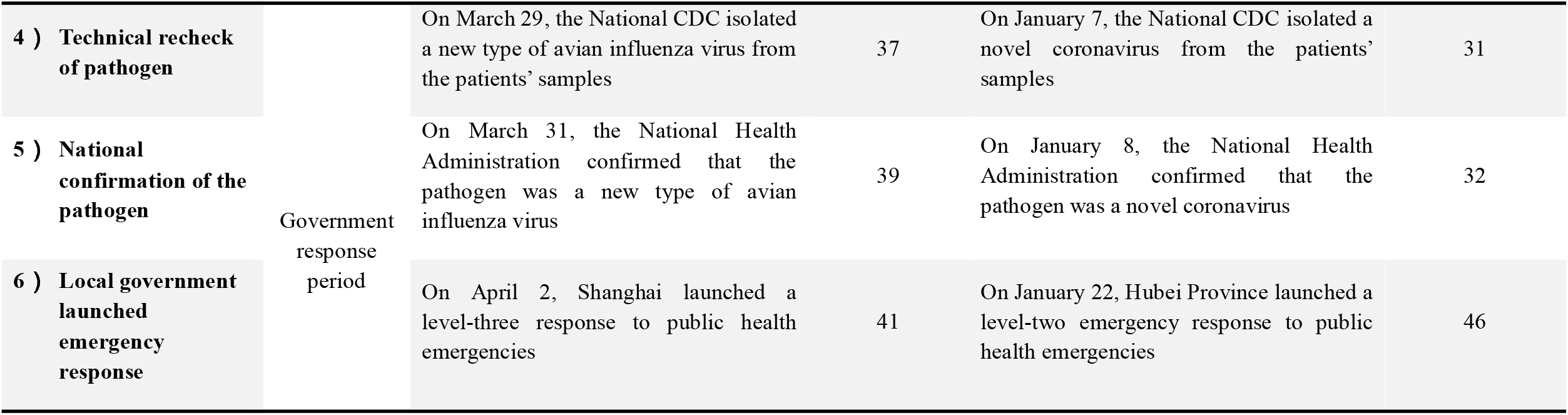
Comparison of the key time nodes of emergency disposal between H7N9 avian influenza (2013) in Shanghai and coronavirus disease 2019 in Wuhan.

**Figure 1.**
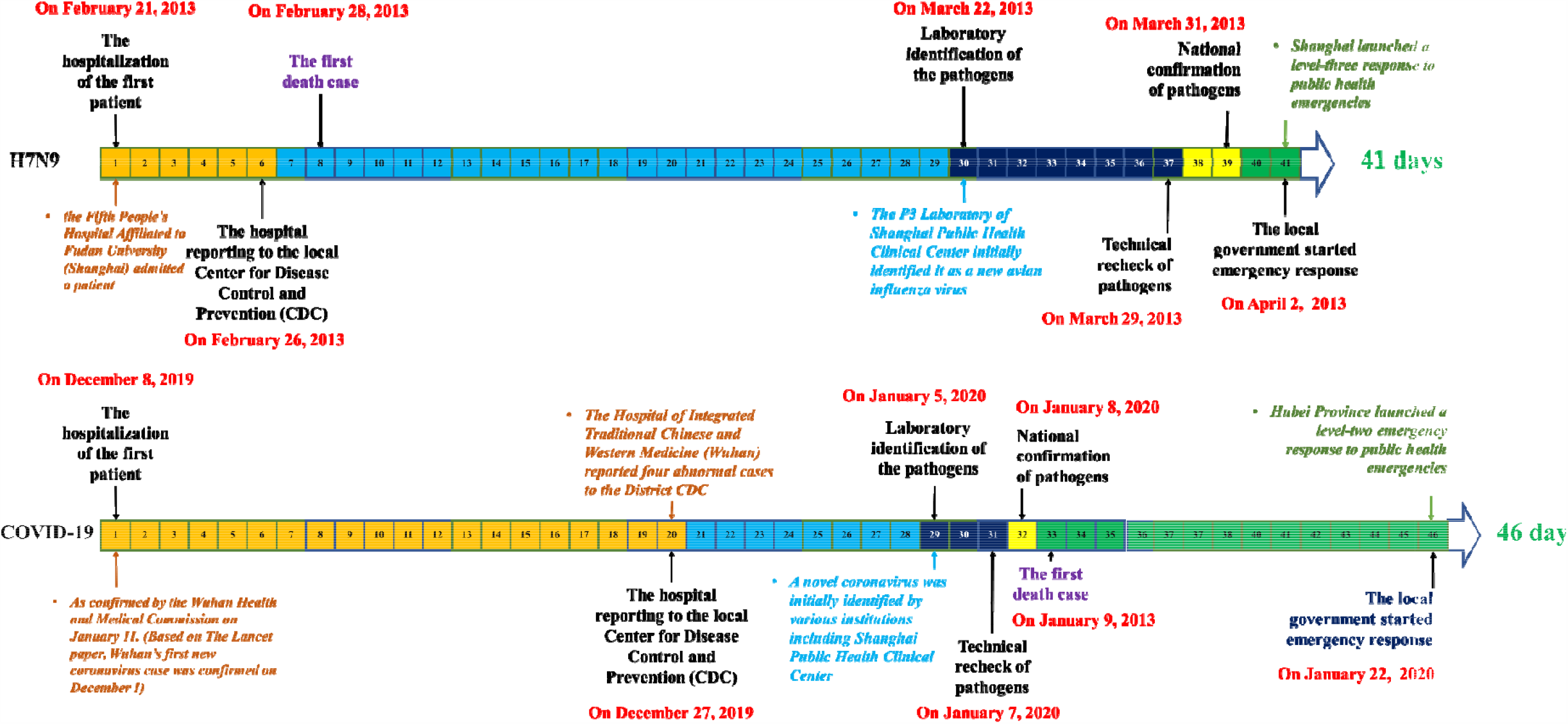
Comparison of the emergency disposal timeline between H7N9 avian influenza (2013) in Shanghai and coronavirus disease 2019 in Wuhan.

### Hospital to CDC reporting period

#### H7N9 avian influenza

The first patient was hospitalized at the Fifth People’s Hospital of Shanghai affiliated to Fudan University on February 21, 2013. Subsequently, two patients were admitted. (16, 17)

*The doctor on duty in the emergency department observed that a paternal relationship existed between the follow-up case and the first case and believed that there was a possibility of transmission. Hence, in the early hours of February 26, 2013 at 1:10 a*.*m*., *he reported his findings to the doctor on duty who was also the chief of the infection department of the said hospital. He believed that the above situation was consistent with the possibility of clustered unexplained pneumonia cases and immediately called the attention of the administrators in charge of the hospital. Subsequently, the hospital gained expert consultation and undertook protective measures. At 2:30 a*.*m. of the same day, the hospital administrators contacted the chief administrative official of the local CDC by telephone and requested the start of epidemiological survey and sampling*. (18)

#### Coronavirus disease 2019 (COVID-19)

The Wuhan Municipal Health Administration announced on January 11, 2020 that the first confirmed case of novel coronavirus pneumonia was detected on December 8, 2019. (18) A literature published in *The Lancet* reported that the first case was detected on December 1, 2019. (19) Based on the principle of caution, this article used December 8, 2019 as the onset time of the first case of the epidemic and considered that this patient was hospitalized at that time.

*On the morning of December 26, 2019, Dr. Jixian Zhang, a doctor from Hubei Hospital of Integrated Traditional Chinese and Western Medicine in Wuhan City, observed an abnormality in a couple’s lung computed tomography (CT) scan and an abnormality in their son’s CT scan as well. Hence, the next day, the hospital reported four abnormal CT findings to the local CDC including another case*. (20)

Hence, the time taken by the hospital to report the first case of H7N9 (2013) in Shanghai and COVID-19 (2019) in Wuhan was 6 and 20 days, respectively.

### Pathogen identification period

#### H7N9 avian influenza

The local CDC conducted an epidemiological survey and sampling at 4:00 a.m. on February 26, 2013 and informed the hospital at 10:30 a.m. that adenovirus, syncytial virus, Legionella, H1N1, highly pathogenic avian influenza virus, Mycoplasma, and seasonal influenza virus tested negative. The hospital subsequently sent the samples to the P3 Laboratory of Shanghai Public Health Clinical Center. On March 22, the Shanghai Public Health Clinical Center preliminarily confirmed the pathogen as a new type of avian influenza virus. On March 29, 2013, the National CDC isolated a new type of avian influenza virus from samples collected from patients.

#### COVID-19

The local CDC was unable to identify the pathogen on December 26, 2019 and subsequently sent the samples to various testing institutions, including Shanghai Public Health Clinical Center and the Chinese Academy of Sciences (Wuhan Virus Institute). Various testing institutions had identified the novel coronavirus and the complete genome sequence between December 30, 2019 and January 5, 2020. (20) On January 7, 2020, the National CDC isolated a new type of coronavirus from the patients’ samples. (22)

Hence, the time taken to identify the pathogen in the cases of H7N9 (2013) in Shanghai and COVID-19 (2019) in Wuhan was 31 and 12 days, respectively.

### Government response period

#### H7N9 avian influenza

On March 31, 2013, the National Health Administration confirmed that the pathogen was a new type of avian influenza virus. On April 2, 2013, the government of Shanghai launched a level-three response to public health emergencies.

#### COVID-19

On January 8, 2020, the National Health Administration confirmed that the pathogen was a novel coronavirus. On January 22, 2020, the government of Hubei Province launched a level-two response to public health emergencies. (22)

Hence, the time taken by the government to respond in the cases of H7N9 (2013) in Shanghai and COVID-19 (2019) in Wuhan City was 4 and 14 days, respectively.

We compared the government’s emergency response process between outbreaks of Shanghai H7N9 avian influenza in 2013 and Wuhan COVID-19 in 2019. The time taken from the detection of the first case to the implementation of public health emergency response was 41 and 46 days for H7N9 avian influenza and COVID-19, respectively. The hospital to CDC reporting period was 14 days slower in the case of COVID-19 than in the case of H7N9 avian influenza. The time taken to identify the pathogen was 19 days faster in the case of COVID-19 than in the case of H7N9 avian influenza. Lastly, the time taken by the government to respond was 10 days slower in the case of COVID-19 than in the case of H7N9 avian influenza (Figure 2).

**Figure 2.**
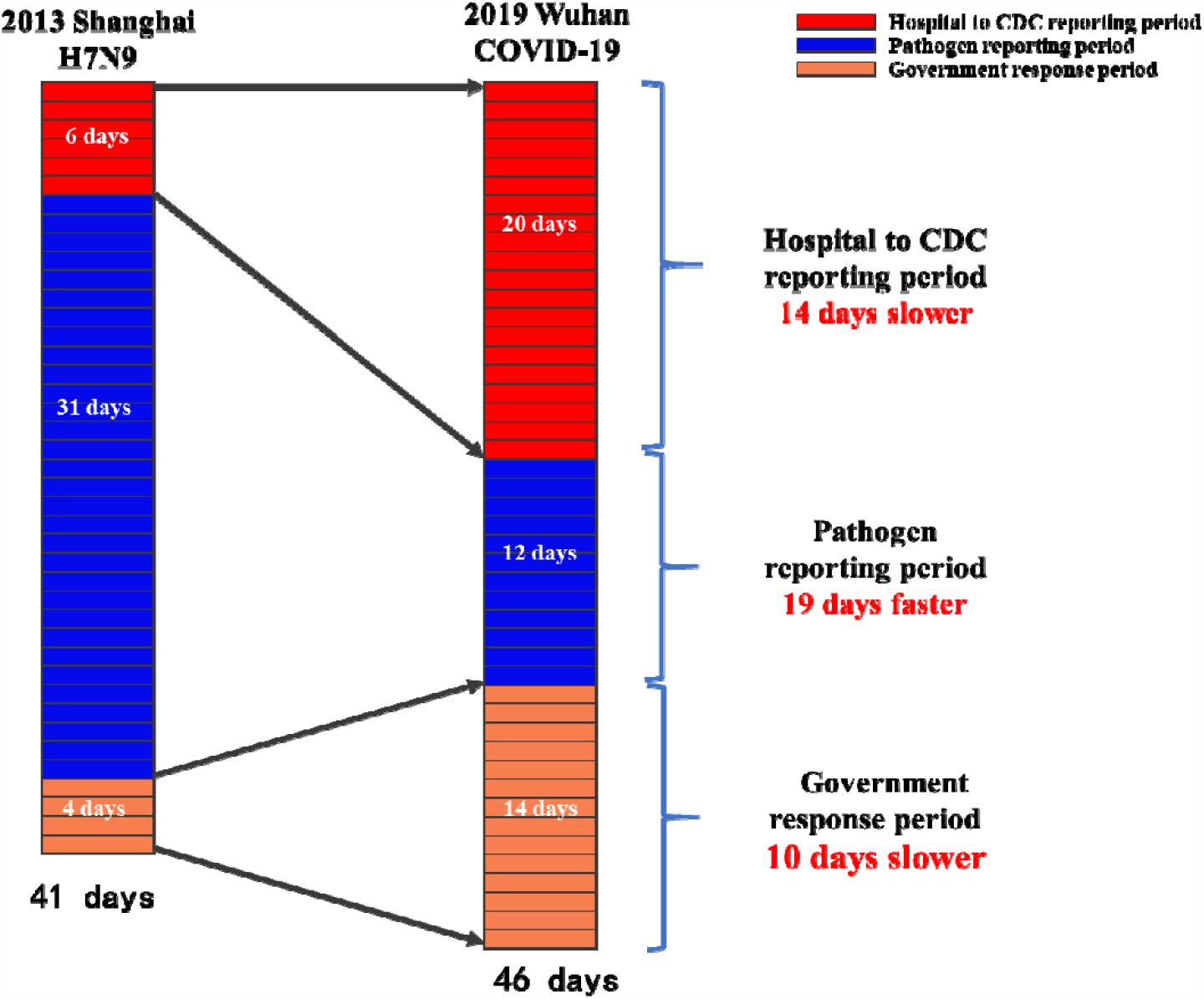
Comparison of three critical emergency disposal speed between H7N9 avian influenza (2013) in Shanghai versus coronavirus disease 2019 in Wuhan.

## Discussion

To the best of our knowledge, this was one of the few studies conducted in China to compare the strengths and weaknesses of public health emergency disposal between COVID-19 and H7N9 avian influenza. In this case-comparative study, the time taken to detect unknown pathogens had improved between the outbreaks of H7N9 avian influenza and COVID-19, whereas the time taken for hospitals to report a case to the local CDC and the government’s emergency response was significantly slow.

In this study, we mainly investigated three crucial periods that influence the efficiency of emergency management to public health crises. During the emergency response process for H7N9 avian influenza (2013) in Shanghai, the maximum time was taken to technically identify and recheck the pathogen. The technical identification of pathogen took 24 days and the rechecking took 7 days, which accounted for 76% of the whole emergency process. In contrast, the time taken to technically identify and recheck the pathogen in the case of COVID-19 was reduced to just 12 days, accounting for 24% of the whole emergency process.

Laboratory identification was 19 days faster in the case of COVID-19 than in the case of H7N9 avian influenza, whereas the total disposal time was 5 days longer in the case of COVID-19 than in the case of H7N9 avian influenza. This could be attributed to the decrease in the reporting periods of certain hospitals and the increase in responding periods of the local governments. The time taken by the hospital to report a case to the local CDC was 14 days slower during COVID-19 than during H7N9 avian influenza (19 days vs. 5 days, respectively). Furthermore, the response period of the local government launching emergency management was 14 days during COVID-19, which was 10 days longer than that during H7N9 avian influenza. Combining the hospital to CDC reporting period and government response period of H7N9 avian influenza with the pathogen identification period of COVID-19 would result in the entire epidemic control taking less than 22 days. Moreover, Hubei Province could thus launch an emergency response on December 30, suggesting that approximately 27 cases of COVID-19 would be detected in Hubei Province and the number of close contacts would be approximately 1350 by early March 2020. The Wuhan Municipal Infectious Diseases Hospital alone had 350 beds, which was sufficient to handle full admission. Subsequently, the local CDC also had sufficient capabilities to screen and isolate most of the patients in close contacts with the infected patients.

The 5-day longer emergency period during COVID-19 could possibly be attributed to the hospital to CDC reporting period and government response period constrained by the following objective conditions:

1. At the beginning stage of the epidemic, H7N9 appeared a larger threat. The duration between the first identified case and the first reported death was only 7 days (on February 28, 2013, the first death case was observed). For COVID-19, this duration was 32 days instead. On January 9, local medical institutions and disease control departments were instructed to speed up and implement isolation and precautionary measures. (20)
2. Because of underreporting of cases considering the challenges in data collection and shortage of testing kits and reagents in Hubei Province. Furthermore, the local medical supplies, beds, and facilities were insufficient, which were even exacerbated by the lockdown of the province, preventing the reach of supplies from several other hospitals.

This study has several potential limitations. First, the assessment coverage was at the city level; thus, comparison between the national level and the grassroot level was not assessed in this study. The grassroot level is the first gateway of public health emergency, and the effective measures and emergency responses taken by the grassroot level are considered important. Second, we used six-time nodes to evaluate the process of the government’s emergency response, which is relatively limited when evaluating the possibility of an epidemic of major infectious diseases. Third, the data are based on China’s official and authoritative reports, coupled with retrospective studies, which inevitably had information bias. Considering all these limitations, the findings should be interpreted with caution before additional studies are conducted.

## Conclusions

The identification of the unknown pathogen has significantly improved in China between the outbreaks of H7N9 avian influenza and COVID-19. However, the time taken for epidemic reports from certain hospitals to reach the local CDC as well as the decision-making process by the local government in Hubei Province was reduced, which might be one of the vital factors for widespread COVID-19 cases. These issues need to be addressed urgently to prepare for public emergencies to prevent and control future epidemics of emerging infectious diseases in China and the world.

## Data Availability

All data, models, and code generated or used during the study appear in the submitted article.

## Contributors

TZ, WS, and LL designed the project, processed and analyzed the data, and wrote the manuscript. YW, GB, RD, and QW edited the manuscript. All authors revised the draft.

## Acknowledgment

This study was supported by the National Natural Science Foundation of China (Nu:71874033) and Key project of Philosophy and Social Science Research of the Ministry of Education (Nu:15JZD029).

